# Advancing Early Warning Systems for Malaria: Progress, Challenges, and Future Directions - A Scoping Review

**DOI:** 10.1101/2024.09.03.24313035

**Authors:** Donnie Mategula, Judy Gichuki, Karen I Barnes, Emanuele Giorgi, Dianne Jannete Terlouw

## Abstract

**Background:** Malaria Early Warning Systems(EWS) are predictive tools that often use climatic and environmental variables to forecast malaria risk and trigger timely interventions. Despite their potential benefits, the development and implementation of malaria EWS face significant challenges and limitations. We reviewed the current evidence on malaria EWS, including their settings, methods, performance, actions, and evaluation.

**Methods:** We conducted a comprehensive literature search using keywords related to EWS and malaria in various databases and registers. We included primary research and programmatic reports focused on developing and implementing Malaria EWS. We extracted and synthesized data on the characteristics, outcomes, and experiences of Malaria EWS.

**Results:** After reviewing 5,535 records, we identified 30 studies from 16 countries that met our inclusion criteria. The studies varied in their transmission settings, from pre-elimination to high burden, and their purposes, ranging from outbreak detection to resource allocation. The studies employed various statistical and machine-learning models to forecast malaria cases, often incorporating environmental covariates such as rainfall and temperature. The most common mode used is the time series model. The performance of the models was assessed using measures such as the Akaike Information Criterion( AIC), Root Mean Square Error (RMSE), and adjusted R squared(R ^2^). The studies reported actions and responses triggered by EWS predictions, such as vector control, case management, and health education. The lack of standardized criteria and methodologies limited the evaluation of EWS impact.

**Conclusions:** This review provides a comprehensive overview of the current status of Malaria EWS, highlighting the progress, challenges, and gaps in the field. The review informs and guides policymakers, researchers, and practitioners in enhancing EWS and malaria control strategies. The review also underscores the need for further research on the integration, sustainability, and evaluation of Malaria EWS usage and harmonized methods to ease review.

## INTRODUCTION

The fight against malaria has reached a critical phase. Despite remarkable strides in reducing the global malaria burden from 2000 to 2015, progress has stalled since 2016, especially in high-burden countries within sub-Saharan Africa [1–3]. This stagnation signals an urgent need for innovative tools and strategies to reinvigorate the push toward the World Health Organization’s (WHO) 2030 targets for malaria elimination[4]. For these elimination targets to not merely be aspirational, there is a need to deploy new or effective vector control measures, diagnostic tools, antimalarial medications and social behaviour change communication. A transformation of malaria surveillance systems is equally important, shifting from passive reporting to dynamic systems capable of tracking hotspots, forecasting outbreaks, and evaluating the effectiveness of interventions [5].

Malaria Early Warning Systems (EWS) are embedded within these enhanced surveillance systems, critical for forecasting and mitigating potential outbreaks. By synthesizing data on intervention strategies, environmental conditions, and resistance patterns, EWS equips health authorities and policymakers with the means to respond effectively to upcoming threats. This proactive approach is vital for reducing the strain on healthcare infrastructure and saving lives. As countries edge closer to malaria elimination, the frequency of outbreaks is expected to increase due to the ‘heterogeneous’ nature of transmission [6,7]. This heterogeneity, driven by disparities in intervention uptake, climate variability, and resistance among vectors and parasites, necessitates robust EWS to navigate and control the evolving landscape of malaria transmission[8]

An effective malaria EWS is a powerful predictive tool, enabling public health officials, governments, and stakeholders to take informed, preemptive actions to prevent impending outbreaks. Integrating data on intervention uptake, climatic conditions, vector and parasite resistance, and other critical factors is essential [9]. However, the implementation and development of EWS face significant challenges. Diverse methodologies to predict future malaria risk present two distinct scenarios: some EWSs provide reasonable certainty (reliability associated with the warning information provided) but inadequate lead time (duration between issuing a warning or alert and the onset of the event) for action. In contrast, others offer good lead time but with modest certainty in predictions. Striking a balance between accurate forecasts and timely response is complex yet crucial in developing effective Malaria EWS [10,11].

The sustainability of investments in malaria EWS is a significant challenge, with funding often being reactive to the occurrence of an outbreak or disaster. This pattern can undermine the long-term effectiveness and maintenance of these systems. For an EWS to be successfully integrated into malaria surveillance, there is a need for improved infrastructure, capacity building, and collaboration among stakeholders, including the community, researchers, and policymakers. These improvements are crucial for the effective functioning and utilization of EWS in malaria prevention and control efforts [12].

The Roll Back Malaria initiative established a framework for malaria EWS in 2001, guiding their development and implementation [13]. However, progress in establishing EWS in Africa has been limited over the past two decades. The first systematic review of malaria EWS, conducted by Zinszer and colleagues in 2012, focused on forecasting methodologies, predictors, and model evaluations but did not address other critical aspects such as actions following early warning predictions, performance evaluation, and integration into existing systems for scalability and sustainability [14]. Subsequent reviews have also been specific in scope [15,16], highlighting the need for a comprehensive and updated review that covers more dimensions of malaria EWS and employs a rigorous, systematic approach to synthesize and analyze the evidence.

This current review aims to summarize the status of malaria EWS, focusing on settings, methods, performance, actions, and evaluation. It seeks to provide insights into the progress, challenges, lessons learned, successes, and limitations of Malaria EWS. The goal is to inform future strategies and enhance the effectiveness of EWS in detecting and responding to malaria outbreaks.

### In this review, we set out to answer the following questions

1. What are the transmission settings and methods used when malaria EWS are developed?
2. What are the performance, usability, and feasibility of the malaria EWS piloted, developed, and implemented?
3. What actions have been documented following malaria EWS predictions?
4. What are the approaches to evaluating a malaria EWS’s performance, effect, and impact?

## METHODS

This review focused on developing and implementing malaria EWS for populations affected by or at risk of malaria, including all age groups and demographics. These systems, either standalone or integrated into broader programs, utilized routinely collected data or data from studies and surveillance systems to predict future malaria risks. Instead of comparing these systems with specific alternatives, as would be the case in a traditional systematic review, we assessed various characteristics of each malaria EWS. The outcomes evaluated included predictions or forecasts of malaria cases, disease, mortality, and anti-malarial drug resistance. We incorporated primary research published in peer-reviewed journals and available online programmatic reports.

### Eligibility criteria

The studies included met the following inclusion and exclusion criteria.

#### Inclusion criteria

- Primary research in a peer-reviewed journal
- Published or available online programmatic reports
- Developing a prediction model for the prediction and or of malaria cases, deaths, drug resistance
- Malaria is the disease of interest.

o Specific outcomes include malaria cases, disease, death, and antimalarial drug resistance.
- Presents the development, evaluation, or other experiences of an EWS in a standalone setting or as part of a program.

#### Exclusion criteria

- Studies focus solely on general malaria trends, risk factors, or predictors without examining or forecasting the specific outcomes related to EWS.
- Studies focusing on malaria in non-human subjects, such as animal or *in vitro* studies.
- Any study that does not explicitly address a prediction model, including the development, evaluation, or other experiences with EWS.
- Studies that do not present predictions or forecasted outcomes for malaria cases, disease, death/mortality, and antimalarial drug resistance.
- Unpublished research, non-peer-reviewed journals

#### Information sources

We used the EBSCOhost platform, which gave us access to several major databases: Medline Complete, Global Health, CNHL Complete, and Green File.

#### Search strategy

Keywords

“malaria” as Medical Subject Headings (MeSH) teams and as a keyword

AND

“early warning” OR “prediction” OR “forecasting” as MeSH terms

And as accessible text terms truncated as follows predict* OR forecast* OR “early warn*“

#### Limiters

- Published between January 2012-30 June 2023( there was a prior review done before 2012[14])
- Online full text available
- Peer reviewed
- Human

### Data management

#### Study selection

The Rayyan tool was used to manage the extracted studies. Two reviewers (DM and JG) independently screened all retrieved studies’ titles and abstracts and excluded irrelevant studies based on eligibility criteria. The full-text screening was conducted for all potentially eligible studies to assess for eligibility based on the inclusion and exclusion criteria. Any disagreements between reviewers were resolved through discussion.

#### Data extraction and management

Data extraction used a pre-designed standard data extraction form developed in-house. This form included vital information such as the author(s), year of publication, study location, study year, study setting, study design, and EWS description parameters.

### Risk of bias assessment

The risk of bias in the included studies was not formally assessed because the scope of this review did not encompass a full systematic approach, which typically necessitates such an evaluation.

However, inclusion and exclusion criteria were strictly applied to maintain the integrity and reliability of the findings.

### Data synthesis

The synthesis of data from the included studies was structured to provide a comprehensive understanding of malaria EWS. This process included a summary of the characteristics of the studies, including the methodology, population, geographic location, and primary findings. A thorough narrative synthesis was done to align with the specific objectives of the review, providing an in-depth analysis and synthesis of key findings. These included: (1) Exploration of the settings and methods used in developing each malaria EWS, data sources, prediction modeling, and technological platforms, (2) Operational aspects of the malaria EWS, focusing on effectiveness, user experience, implementation challenges, and sustainability in diverse contexts, (3) Actions and responses triggered by EWS predictions with identification of successful practices, challenges, and coordination among various stakeholders, and (4) Review of methodologies and criteria used to evaluate EWS’ performance, effect, and impact.

## RESULTS

Following our search, we retrieved 5,535 records from the databases (Table 1). No additional records were identified from the programmatic reports registers. After 980 duplicate records were removed, titles and abstracts of the remaining 5,535 records were screened. After excluding ineligible articles, we identified 57 studies for a detailed full-text evaluation of their eligibility. During full-text review, we excluded 27 that did not meet the inclusion criteria. The narrative provided in this review is derived from a synthesis of 30 articles (Figure 1).

**Figure 1.**
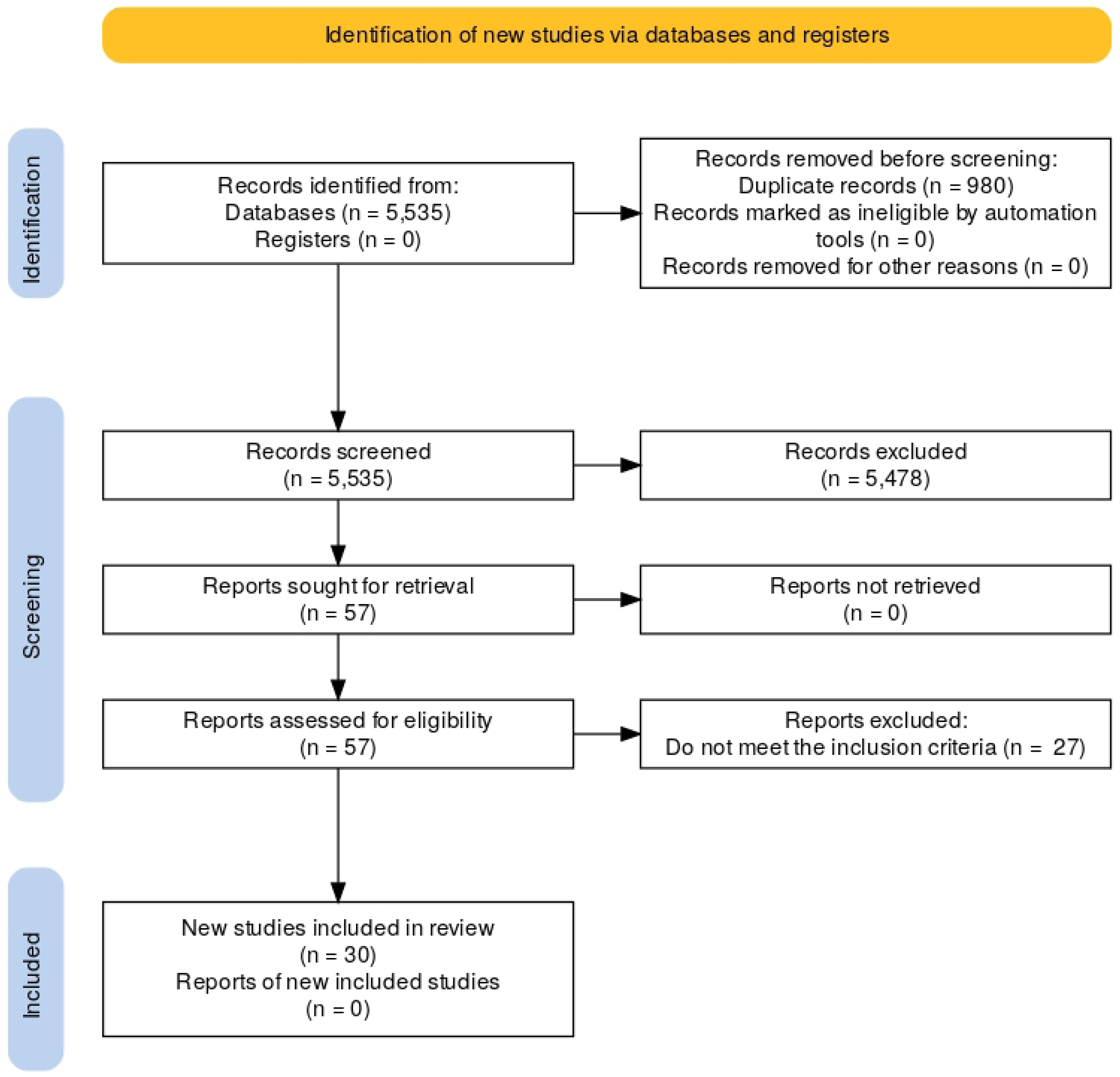
Review Flow diagram.

**Table 1.**
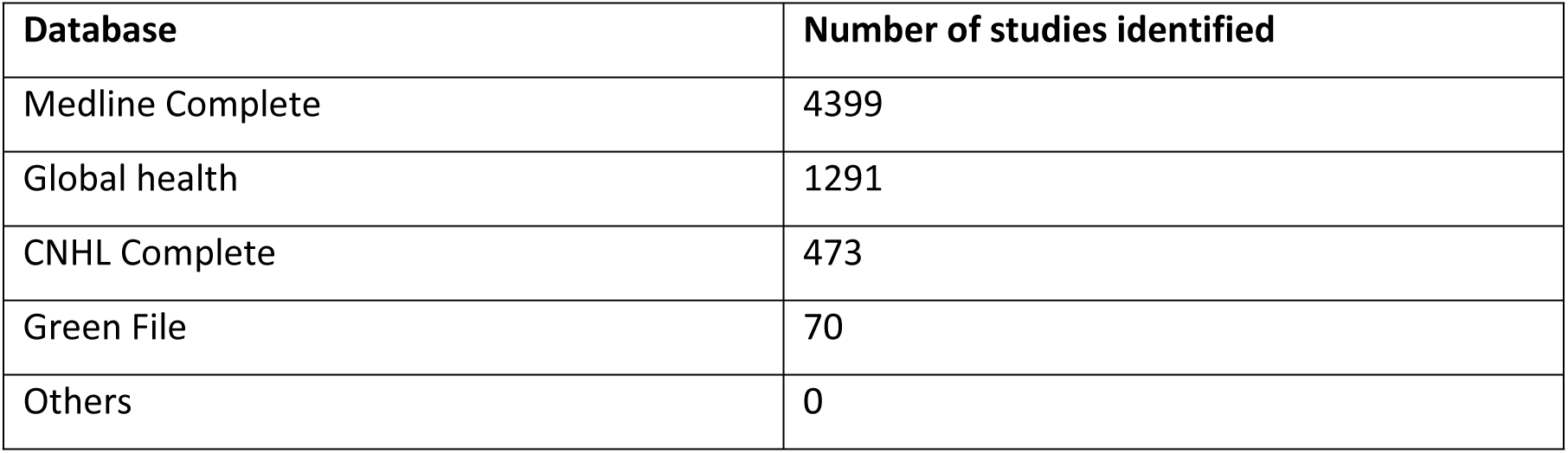
Number of studies identified through each database search (including duplicates)

| Database | Number of studies identified |
| --- | --- |
| Medline Complete | 4399 |
| Global health | 1291 |
| CNHL Complete | 473 |
| Green File | 70 |
| Others | 0 |

All data collated can be found in supplementary file 1.The review identified studies from 18 countries, with India contributing the largest proportion and accounting for seven studies in total. South Africa and Kenya followed, as illustrated in Figure 2 below.

**Figure 2.**
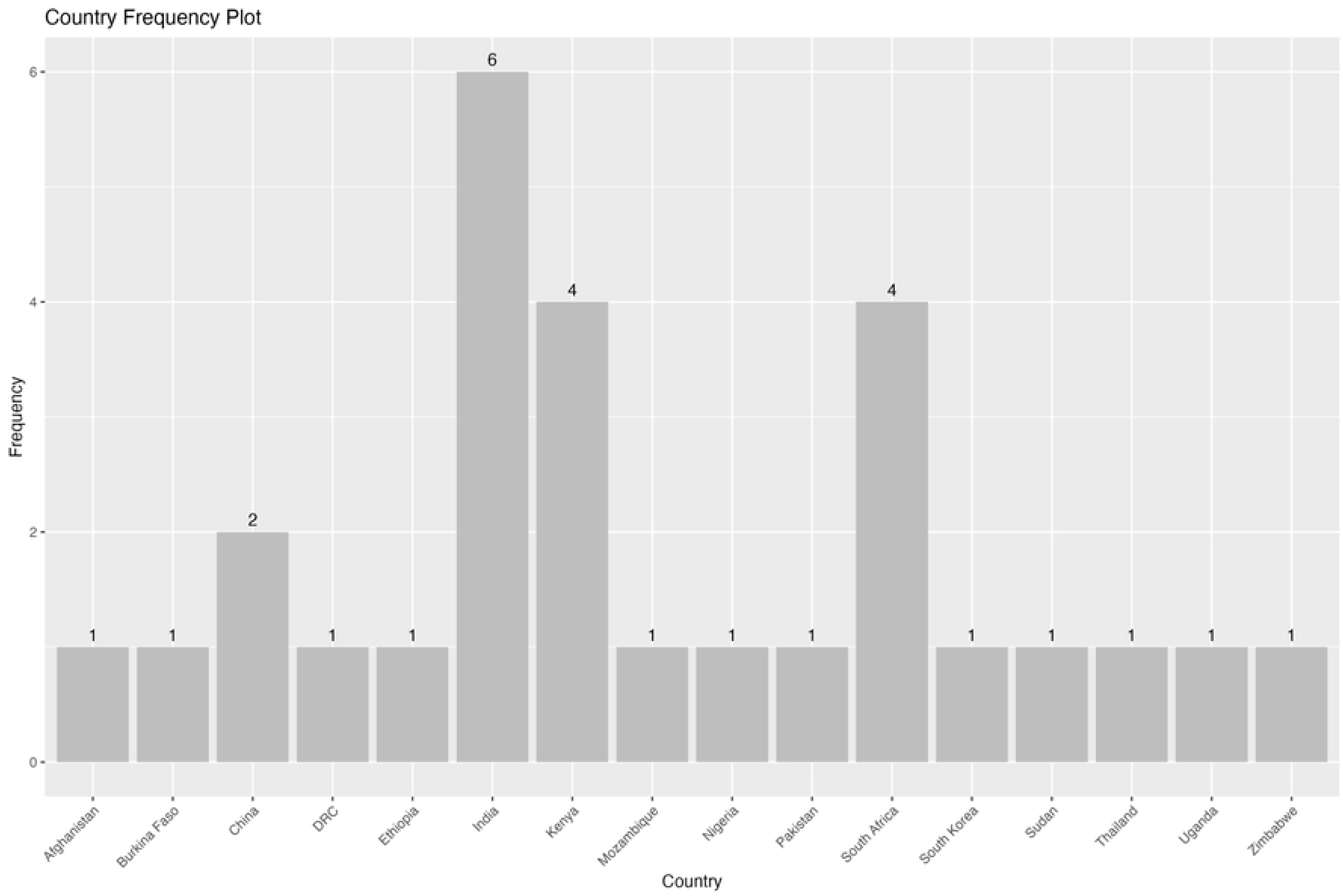
Frequency plot showing the countries where included studies were conducted.

### Settings where malaria EWS were developed

Our systematic review identified a range of papers on Malaria EWS across varied transmission settings. These settings include seasonal transmission areas such as Afghanistan [17] and Pakistan[18], where EWS models predict malaria trends based on climatic fluctuations. We also reviewed papers from highly heterogeneous regions like Ethiopia[19] and Kenya [22], where EWS focuses on a high transimmision setting of the country. Perennial transmission settings are represented by several studies from India[20–26] and other areas[27,28], demonstrating year-round prediction systems. Some malaria EWS papers covered regions nearing malaria elimination, such as South Africa [29–32], focusing on detecting residual transmission [33]. Other aspects included regions with high transmission during rainy seasons [34], provinces with malaria epidemics in China[35], areas with stable transmission [36] and those with low risk [37]. Some studies focused on specific challenges, such as the re-emergence of malaria in border regions in China[38], and leveraging indigenous knowledge in high-risk zones in Zimbabwe [39](Figure 3).

**Figure 3.**
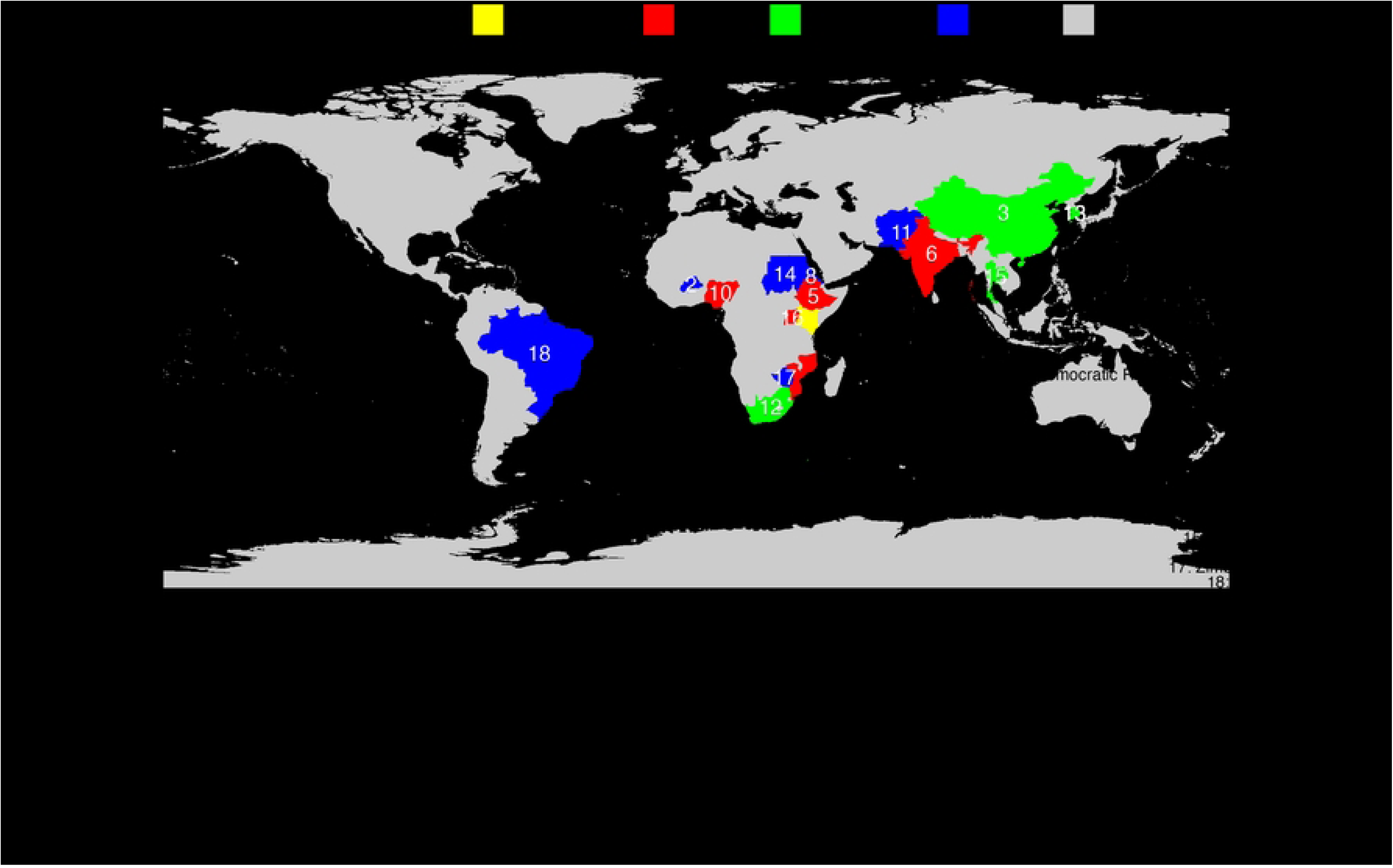
Countries where the included studies were done and their malaria transmission setting.

### Approaches to Developing EWS

Several approaches were used to develop malaria EWS (Table 2). We broadly classified the models into statistical models, machine learning models, and indigenous knowledge. Statistical approaches typically rely on predefined models where covariates are carefully selected based on prior knowledge and are explicitly included in the model structure to explain the relationship between variables. These models often assume a specific distribution for the data and emphasize interpretability, allowing for clear inferences about the effects of each covariate. In contrast, machine learning approaches often focus on predictive performance rather than interpretability, using algorithms that can automatically select, transform, and weigh covariates in complex ways without requiring prior assumptions about the data’s distribution. While statistical models typically convey uncertainty through confidence intervals and p-values, machine learning models often rely on techniques such as cross-validation and bootstrapping to estimate uncertainty, though these estimates may not always be directly interpretable.

**Table 2.** Approaches to the development of EWS.

| **Approach Category** | **Methods Used** | **Studies/References** |
| --- | --- | --- |
| Statistical | Correlation | Dhiman (2017) |
| Statistical | Dynamic System | Harris (2020) |
| Statistical | Geostatistical model | Colborn (2018) |
| Statistical | Process | Roy (2015) |
| Statistical | Regression | Bouma (2016), Sewe (2017), Verma (2018) |
| Statistical | Time series | Adeola (2014), Anwar (2015), Ebhuoma (2016), Hussien (2017), Karuri (2018), Kifle (2019), Kumar (2020), Mopuri (2023), Riaz (2023), Wang (2023), Zinszer (2023) |
| Statistical | Time series, Multimodel | Panzi (2022) |
| Statistical | Time series, non linear | Kim YoonHee (2019), Ratnam (2019) |
| Machine Learning | General Machine Learning | Mohapatra (2021) |
| Machine Learning | Neural Networks | Barboza (2016), HADDAWY (2020), Kamana (2022), Santosh (2022) |
| Machine Learning | Rule based | Buczak (2015) |
| Machine Learning | Supervised Learning | Harvey (2021) |
| Indigenous Knowledge | Community EWS | Macherera (2016) |

Most of the studies (12) used statistical models, primarily using time series analysis such as AutoRegressive Integrated Moving Average (ARIMA) [17,21,40]. Some studies used variations of ARIMA, such as Seasonal AutoRegressive Integrated Moving Average (SARIMA) and Seasonal AutoRegressive Integrated Moving Average with Exogenous Regressors (SARIMAX), to account for seasonal adjustments and exogenous variables[23,36] . As demonstrated in the paper by Panzi and colleagues, Multimodal approaches were employed to construct malaria EWS models integrating various statistical methods[41].

Other studies utilized regression models to quantify the relationship between variables[42] or correlation models to identify the strength of associations[20]. Some studies applied dynamic systems theories and process models to predict malaria trends. For instance, dynamic systems theories, such as the theory of critical slowing down[43], were used to anticipate shifts in malaria transmission under varying conditions. Similarly, process-based models [24] were employed to simulate the complex interactions between environmental factors and malaria dynamics, providing insights into how the disease may evolve.

We identified nine studies that utilized machine learning models for EWS development. Studies done by Harvey and colleagues and by Martineau and colleagues applied supervised learning methods, including Gaussian Processes and Random Forests, to predict malaria epidemics[32,34]. Neural networks, particularly Long Short-Term Memory (LSTM) models, were used in four studies by Kamana Santosh, Barboza, and Haddway for their effectiveness in handling large datasets and complex patterns, such as climate change effects and city-specific malaria trends[25,38,44].

Additional approaches included the use of the Waikato Environment for Knowledge Analysis (WEKA) by Mohapatra et al. [22] for classifier selection and a combination of Generalized Linear Models (GLM), Ensemble Methods (EM), and Support Vector Machines (SVM) by Brown and colleagues [27]for data-driven predictions in dense populations. Techniques like fuzzy association rule mining by Buczak et al. [37] and Bayesian networks by Haddway and colleagues [33] were also used. One study, the Gwanda District study in Zimbabwe[39], utilized Indigenous Knowledge Systems for their malaria EWS.

### Conveying uncertainty of predictions

Seven studies incorporated measures of uncertainty in their predictions, primarily using confidence intervals. Anwar and colleagues [17] predicted malaria cases in Afghanistan from January 2014 to September 2015, providing confidence intervals to express the uncertainty in their forecasts. Similarly, Roy [27], Karuri [33], Ebhuoma [39], and Zinszer [47] included confidence intervals in their predictions. Panzi [44] also used confidence intervals in forecasting malaria cases in the Democratic Republic of the Congo (DRC) from 2020 to 2030. In contrast, Colborn [31] employed non-exceedance probabilities, an alternative method for representing predictive uncertainty. This distinction highlights the different approaches to quantifying and communicating uncertainty across these studies. The models that did not include uncertainty are mostly used machine-learning models

### Covariates and Data Sources

A range of covariates was used in the included studies (Table 3). Standard covariates across the studies include environmental factors such as rainfall, temperature, and humidity, alongside vegetation indices like normalized difference vegetation index (NDVI) that measures vegetation health and density and enhanced vegetation index (EVI), which is helpful in areas with dense vegetation. Some studies have considered demographic data and specific climate indices like the Oceanic Niño Index (ONI). A few studies did not include covariates, relying solely on malaria case reports. Additionally, the identified studies have utilized various data sources, including public health reports, national disease control databases, health facility data, and satellite-derived data.

**Table 3.** The covariates included in the models and data sources.

| **Year** | **Author** | **Covariates** | **Data Source** |
| --- | --- | --- | --- |
| 2016 | Anwar | Rainfall, NDVI, EVI, NDWI | Ministry of Public Health Reports (2005-2015) |
| 2021 | Harvey | Not included | Integrated e-Diagnostic Approach (leDA) Database |
| 2022 | Kamana | Temperature | Chinese Centre for Disease Control and Prevention |
| 2019 | Wang | Temperature, humidity, air pressure, vapor pressure, moisture level, wind velocity, precipitation, sunshine duration, days with daily precipitation | Yunnan Province Malaria Data (2011-2017) |
| 2022 | Panzi | Rainfall, temperature, humidity, wind speed | DRC Epidemiological Surveillance Directorate Database |
| 2016 | Bouma | Sea surface temperature over the Pacific and Indian Oceans | Malaria Case Reports (1982-2005) |
| 2017 | Dhiman | Monthly Oceanic Nino Index (ONI) | CBHI & NVBDCP Data (1994-2015) |
| 2014 | Kumar | Rainfall, temperature, wind Speed, humidity | Public Health Data (2006-2013) |
| 2021 | Mohapatra | Rainfall, temperature, humidity, topography | Directorate of Public Health Services, Odisha Data (2002-2017) |
| 2020 | Mopuri | Rainfall, temperature, NDVI | NVBDCP, Visakhapatnam Data (2001-2016) |
| 2015 | Roy | Rainfall, population | National Institute of Malaria Research Data |
| 2020 | Santosh | Temperature, rainfall, age, sex, vegetation index | RoutineEpidemiological Data (1995-2018) |
| 2018 | Verma | Not included | Data obtained via Google Search |
| 2020 | Harris | Not included | Hospital Case Reports (1965-2002) |
| 2016 | Karuri | Rainfall | Pediatric Malaria Admission Data (1990-2011) |
| 2019 | Kifle | Rainfall | Malaria Incidence Data (2012-2016) |
| 2017 | Sewe | LSTs (Day and night), precipitation | Siaya District Hospital Admission Data (2003-2013) |
| 2018 | Colborn | Not included | NMCP Routine Data |
| 2020 | Brown | Demographics, temperature, rainfall | Hospital Routine Data |
| 2023 | Riaz | Not included | MOH Routine Data |
| 2019 | Adeola | Rainfall, NDVI, EVI, NDWI | Malaria Case Observations (2013-2017) |
| 2018 | Ebhuoma | Rainfall, Temperature, Wind Speed, Humidity | Health Facility & Satellite-Derived Data |
| 2019 | Kim YoonHee and Ratnam | Temperature, precipitation | Malaria Case Data in Vhembe, Limpopo (1998-2015) |
| 2022 | Martineau | Sea surface temperature over the Pacific and Indian Oceans | Malaria Institute, Tzaneen Data (1998-2020) |
| 2017 | Hussien | None used | Routine Incidence Data |
| 2016 | Haddway | Environmental variables, time lagged effects | Community Reports & Environmental Data |
| 2015 | Zinszer | Rainfall, temperature, vegetation, clinical data | Health Facility & Satellite-Derived Data |
| 2016 | Macherera | Insects, plant phenology, animals, weather, cosmological indicators | Community Reports & Indigenous Environmental Indicators |
| 2022 | Barboza | Not included | Not included |
| 2015 | Buczak | Rainfal, Tempereture , social data, intervention data | Korea Centers for Disease Control and Prevention website for the period from 2004–2013. |

NDVI: Normalized Difference Vegetation Index EVI: Enhanced Vegetation Index NDWI: Normalized Difference Water Index leDA: Integrated e-Diagnostic Approach DRC: Democratic Republic of the Congo LST: Land Surface Temperature CBHI: Community-Based Health Insurance NVBDCP: National Vector Borne Disease Control Programme NMCP: National Malaria Control Program MOH: Ministry of Health

**Table 4.**
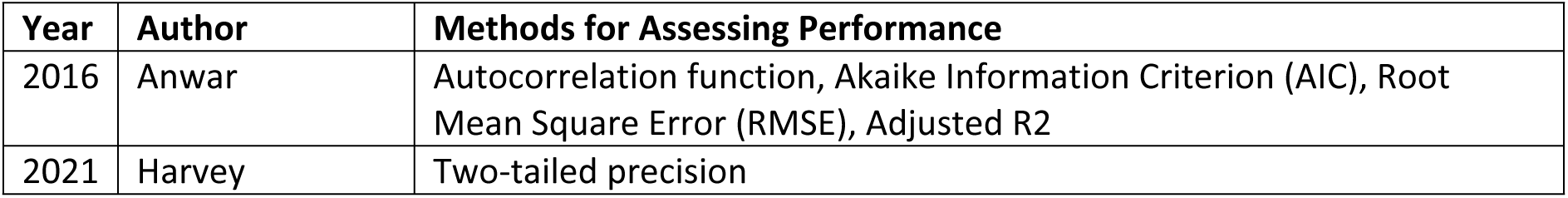
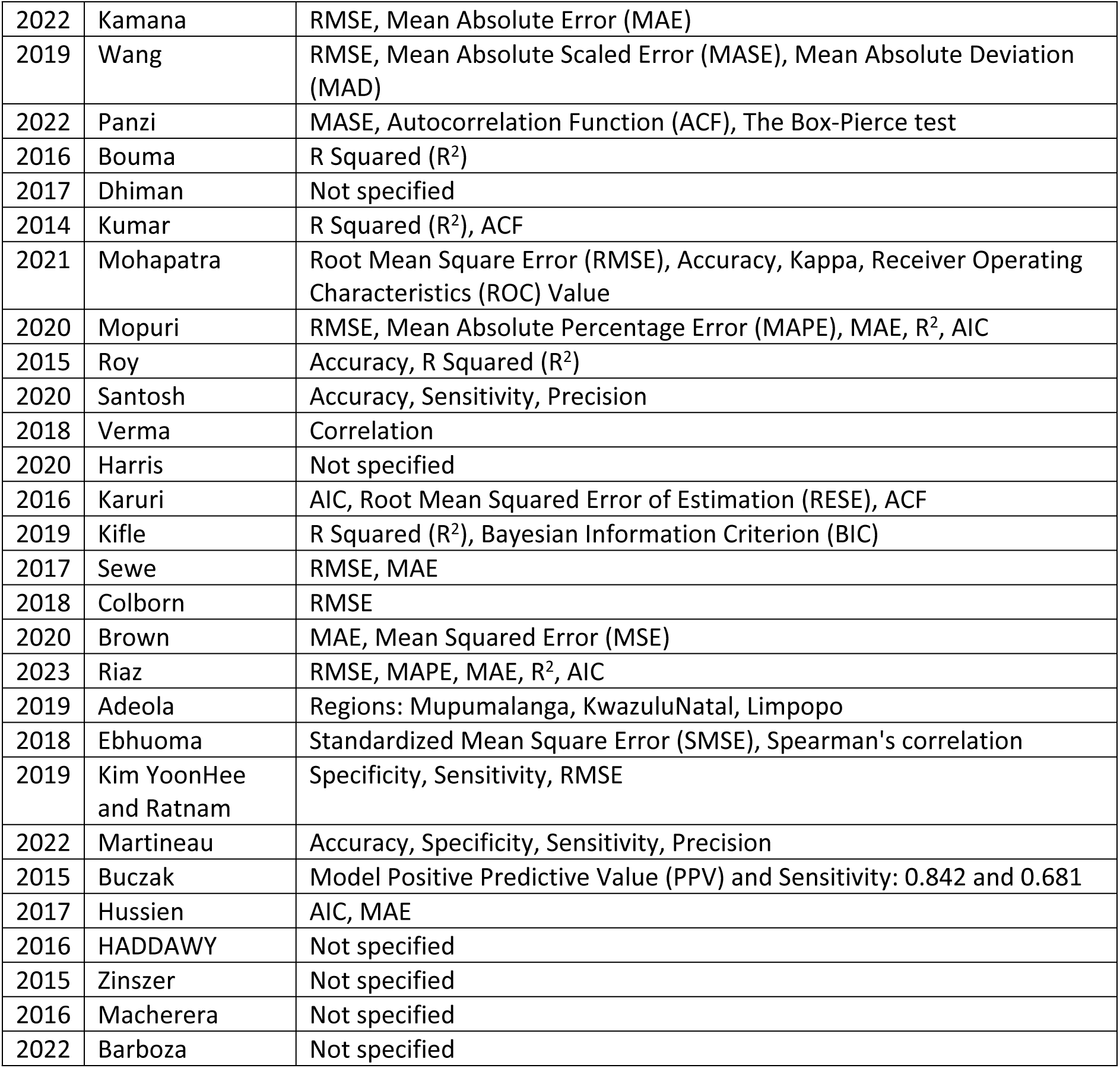
Assessing the performance of the EWS models.

### Assessments of EWS model performance

Table 5 below summarises the EWS assessment methods used. Commonly used metrics for assessing performance were the Root Mean Square Error (RMSE) , [18,31,35,38], Mean Absolute Error (MAE) [23,27,38,42] and R Squared (R^2^) value [19,21,24], alongside more complex statistical tools like the Akaike Information Criterion (AIC)[17,18,23] and Bayesian Information Criterion (BIC)[45]. Some studies have focused on precision measures such as accuracy, sensitivity, and specificity. In contrast, others have utilized auto-correlation functions and error estimation methods. In general, the models provided good predictability based on the methods used.

### Outbreak detection

Outbreak detection methodologies varied across the studies. Harvey and colleagues defined a malaria outbreak occurrence as the point at which the case rate surpassed the five-year mean for the same period plus two standard deviations, providing a statistically significant signal of an outbreak[34]. Roy and colleagues [24] utilized a binary classifier to predict large outbreaks, defining an outbreak occurrence as the point when the probability exceeded a set threshold, which was then validated against actual data. Harris and colleagues determined outbreaks by a substantial increase in cases, specifically when the count exceeded the previous months’ numbers by more than two and a half times [43]Lastly, Colborn and colleagues used exceedance probabilities (EPs) of relative risk to define outbreak thresholds, offering a probabilistic approach to outbreak detection [46].

### Actions following early warning predictions and incorporation into routine practice

In two studies, malaria EWS development went beyond development, and a report was included on incorporating the EWS into routine practice. For instance, they successfully integrated their malaria EWS into the district-level routine practice, streamlining the outbreak detection and response process, including the distribution of bed nets, indoor residual spraying, and larviciding [34]. Similarly, the EWS approach by Macherela at the ward level demonstrates the system’s adaptability and effectiveness in local settings, improved community awareness, and facilitated education campaigns [39].

## DISCUSSION

The importance of malaria forecasting within the public health area cannot be overstated. The origins of malaria EWS can be traced to rudimentary forecasting methods pioneered by health practitioners in the 1900s who forecasted malaria using weather data[48]. These have evolved into more sophisticated models to support control and elimination efforts[14]. Our review identified 5,535 records, including 30 studies that have enriched our understanding of malaria early-warning systems. Through these findings, we lay the groundwork for assessing the current practices and gaps within malaria forecasting and EWS development.

Forecasting methods should be scrutinized for their underlying assumptions, strengths, and weaknesses, with accuracy evaluations conducted on out-of-sample data. We acknowledge numerous forecasting methods, but it is of value to leverage standard forecasting measures to enable cross-study comparisons. Our review found that time series forecasting methods, particularly regression-based approaches, are advantageous due to their flexibility and intuitive appeal. However, their limitations include a tendency to overlook serial autocorrelation in errors, potentially leading to bias in predictor effects and underestimated standard errors. Such models’ residuals should be examined for autocorrelation[49,50]. We also identified studies that use ARIMA models, which can manage serial autocorrelation in the data, with their extended variants like SARIMA and ARIMAX providing additional predictive and forecast capabilities. However, these models require a substantial amount of data and examination of residuals to avoid misleading cross-correlation functions, which still need to be manually done in some cases[51]. Other complex methods in the machine-learning space have also been used. Studies reviewing the use of machine learning models in malaria EWS show their versatility across various ecosystems and capability to achieve greater accuracy but point to the need for their standardization to allow for assessment across models[52] The current landscape of malaria forecasting is quite strong, mainly due to a solid foundation in methodology. Looking ahead, the focus should shift towards enhancing the performance of these models, refining their user interface, and automating their functions to facilitate their adoption by stakeholders within malaria-affected countries. To this end, it is essential to prioritize the development of intuitive platforms that can be seamlessly integrated into existing health systems’ workflows. An example is the EPIDEMIA system used in the Amhara region of Ethiopia, which utilizes near-real-time environmental data and patient records to provide updated malaria risk maps and forecasts. Such systems allow health officials to make timely decisions and improve intervention strategies based on current data rather than historical trends[53,54]

There is also a clear need to streamline these models to operate with (near) real-time data, enabling dynamic responses to evolving malaria trends. This could include developing adaptive algorithms that learn and improve from each prediction cycle, thereby increasing the accuracy and reliability of the forecasts. We also think fostering open-source communities around these models can accelerate innovation, allowing for collective problem-solving and sharing of best practices[55].

Another critical area is customizing these models to account for local environmental variables, socio-economic factors, and intervention strategies, which are crucial determinants of malaria transmission. As we move forward, it is also essential to consider the scalability of these models, ensuring they can be deployed in various settings, from rural clinics to national public health centers. We must also invest in capacity building, providing training and support to local health practitioners and decision-makers to leverage these tools effectively. This approach will increase the reach of malaria EWS and empower local actors to take charge of malaria mitigation efforts in their communities[56].

In addition to refining existing malaria forecasting models, there is an urgent need to expand the EWS scope to predict not just the incidence of malaria but also critical outcomes like mortality rates and the emerging threats of antimalarial drug or insecticide resistance [57]. We found this to be a crucial gap in our review. The capacity to forecast these outcomes would be a significant leap forward, enabling health systems to allocate resources for immediate case management and long-term strategic planning. Predictive models that can, for example, anticipate the spread of drug-resistant strains could inform more effective malaria treatment policy decisions and guide research into new treatments. This broadening of focus will ensure that forecasting models remain relevant and potent tools in the evolving landscape of malaria control and prevention efforts.

The review found few documented actions following early warning predictions for malaria. These actions could range from mobilizing public health resources and the preemptive distribution of anti-malarial medications and bed nets to targeted vector control measures such as indoor residual spraying and larviciding. They could also be community awareness and education campaigns often intensified to improve prevention and early treatment-seeking behaviors.[34,39]

Despite the recognized importance of Malaria EWS, the literature needs to be more extensive in terms of their evaluations of their performance, effect, and impact after implementation. Such evaluations are necessary to ascertain the true efficacy of these systems. With assessment protocols and outcome data, refining EWS, tailoring them to specific environments, and justifying their adoption within routine health practices becomes easier. It is also essential to establish a standardized operational definition of an outbreak[58]. The diverse definitions used across studies make it challenging to compare methods or determine which is more effective. This variability arises from the different approaches to defining an outbreak, whether it be a specific threshold of cases, expert judgment, or complex data models. Without a standard definition, each study may define an outbreak differently, leading to inconsistencies in model evaluation and interpretation[58]

## CONCLUSION

For malaria forecasts to be actionable in public health and clinical settings, they must be accurate (while acknowledging uncertainty), have appropriate spatial and temporal resolution, and consider the operational context, including the availability of data and the technical skill required for model application. Using different forecasting methods on identical datasets, coupled with exploring a broader array of predictors, including transmission-reducing interventions and standardized forecast accuracy measures, will assist in refining malaria forecasting models. The future of malaria forecasting hinges on making the existing models more precise, user-friendly, and automated while ensuring they are adaptable, scalable, and accessible to health professionals across the spectrum of malaria-affected regions. Through these initiatives, we can enhance the global malaria response and move closer to the ultimate goal of malaria elimination

## Data Availability

The datasets analyzed during the current study are available from the corresponding authors of the original studies upon reasonable request.

